# Cost-effectiveness of sotagliflozin for the treatment of patients with diabetes and recent worsening heart failure

**DOI:** 10.1101/2023.12.05.23299552

**Authors:** Jaehong Kim, Shanshan Wang, Slaven Sikirica, Jason Shafrin

## Abstract

**Background:** The efficacy and safety of sotagliflozin, a novel therapy for the treatment of patients with heart failure, was demonstrated in SOLOIST-WHF, but its economic value is yet to be determined. This study assesses the cost-effectiveness of sotagliflozin for the treatment of patients hospitalized with heart failure and comorbid diabetes.

**Methods:** An economic model with a Markov structure was created for patients hospitalized for heart failure with comorbid diabetes. Outcomes of interest included hospital readmissions, emergency department visits, and all-cause mortality measured over a 30-year time horizon. Baseline event frequencies were derived from published real-world data studies; sotagliflozin’s efficacy was estimated from SOLOIST-WHF. Health benefits were synthesized using quality-adjusted life years. Costs included pharmaceutical costs, rehospitalization, emergency room visits and adverse events. Economic value was measured using the incremental cost effectiveness ratio.

**Results:** Sotagliflozin use decreased annualized rehospitalization rates by 34.5% (0.228 vs. 0.348, difference: -0.120), annualized emergency department visits by 40.0% (0.091 vs. 0.153, difference: -0.061), and annualized mortality by 18.0% (0.298 vs. 0.363, difference: -0.065) relative to standard of care, resulting in a net gain in quality adjusted life-years of 0.425 for sotagliflozin vs. standard of care. Incremental costs using sotagliflozin increased by $19,374 over the lifetime of the patient, driven largely by increased pharmaceutical cost. Estimated incremental cost effectiveness ratio was $45,596 per quality adjusted life-year.

**Conclusions:** Sotagliflozin is a cost-effective addition to standard of care for patients hospitalized with heart failure and comorbid diabetes.

## Introduction

The prevalence of heart failure in the U.S. is estimated to be 6.7 million, or about 2.3%, causing approximately 84,000 deaths annually ^1^. Patients with heart failure often suffer from multiple comorbidities and chronic conditions such as type 2 diabetes ^2^. In fact, among patients hospitalized for heart failure, 44% are also diagnosed with type 2 diabetes, a proportion significantly higher than the 10 to 15% of the general population that are diagnosed with diabetes alone ^3^. To care for patients with heart failure, clinical guidelines recommend use of the four classes of medical therapy for heart failure, which incorporates angiotensin-converting enzyme inhibitor, angiotensin receptor blocker, angiotensin receptor-neprilysin inhibitors, beta-blockers, mineralocorticoid receptor antagonists, and, most recently, sodium-glucose co-transporter type 2 inhibitors (SGLT2) into a comprehensive treatment program; this approach can extend the life of a typical patient aged 65 years by an additional five years compared to a conventional therapy and strategy ^4^.

Sotagliflozin, is an orally delivered, small molecule SGLT1 and SGLT2 inhibitor. Findings from the SOLOIST-WHF trial (NCT03521934), a Phase 3, double-blind, randomized, placebo-controlled trial conducted to understand the safety and potential efficacy of sotagliflozin, demonstrated that the rate of the composite of cardiovascular death and hospitalizations and urgent visits for heart failure was lower in the sotagliflozin group than in the placebo group ^5^. Additionally, sotagliflozin showed efficacy in terms of improved cardiovascular outcomes for patients with heart failure, type 2 diabetes, and chronic kidney disease in the SCORED (NCT03315143) trial ^6^.

This study aimed to measure the cost-effectiveness of sotagliflozin for heart failure treatment among patients with worsening heart failure as well as comorbid type 2 diabetes as studied in the SOLOIST-WHF trial. While other studies have examined the cost-effectiveness of SGLT2 inhibitors, this is the first published cost-effectiveness analysis (CEA) model for a treatment such as sotagliflozin which inhibits both SGLT1 and SGLT2 pathways ^7,8^. For this study, which is limited to the population of patients with type 2 diabetes who had recent worsening of heart failure defined as a recent hospitalization or urgent care visit, we estimated the cost and health outcomes associated with sotagliflozin compared to the standard of care (SoC). Health outcomes include quality-adjusted life years, life years (LYs) gained, hospital readmissions, and emergency department visits, while costs include pharmaceutical and medical costs. The primary outcome of interest was the incremental cost-effectiveness ratio, which represents opportunity costs of additional 1 year with perfect health, and outcomes for the model are measured from a U.S. payer perspective. The health-related quality of life and cost were discounted at 3% annually.

## Methods

### Patient Population

The study population aims to replicate the trial population of the SOLOIST-WHF trial. In that trial, patients were required to be an adult (aged 18 to 85 years) who had been hospitalized or had been treated in an urgent care setting because of the presence of signs and symptoms of heart failure and had received treatment with intravenous diuretic therapy. Patients were also required to have received a previous diagnosis of type 2 diabetes before the index admission or to have laboratory evidence to support a diagnosis of type 2 diabetes during the index admission. This model itself uses a hypothetical cohort of adults who were hospitalized for heart failure comorbid with type 2 diabetes.

### Model Overview

The model structure relied on a first-order Markov chain with a one-month cycle length and a 30-year time horizon and was simulated using a hypothetical cohort of 1,000 hospitalized heart failure patients who were discharged from hospitals last month and have been stabilized following hospitalization for heart failure (Figure 1). The approach largely followed the model structure of previous heart failure CEA, which largely employ Markov models with health states of stable disease, hospitalization, and death ^7,9–12^. Our model built on these frameworks but included two key additions. First, we included not only stable disease, hospitalization, and death health states, but emergency department visits as well as emergency department visit rates are not negligible and have remained stable overtime^5,13^. Second, the model allowed for differential hospital readmission rates depending on the time from discharge to capture how the risk of readmission changes over time for heart failure patients. Specifically, the model structure dynamics allowed for a patient in stable disease to have different probabilities of rehospitalizations by 1-30 days, 31-60 days, 61-90 days, or more than 90 days from the last discharge. Empirically, it is well known that readmission rates in the period 1-30 days are higher than those 31-60 days after discharge and much more than 61-90 or >90 days after discharge ^14,15^. To incorporate this empirical finding, we assumed that each study cycle of 1 month, a patient is readmitted to a hospital with a rate of rehospitalization associated with days from the last discharge.

**Figure 1:**
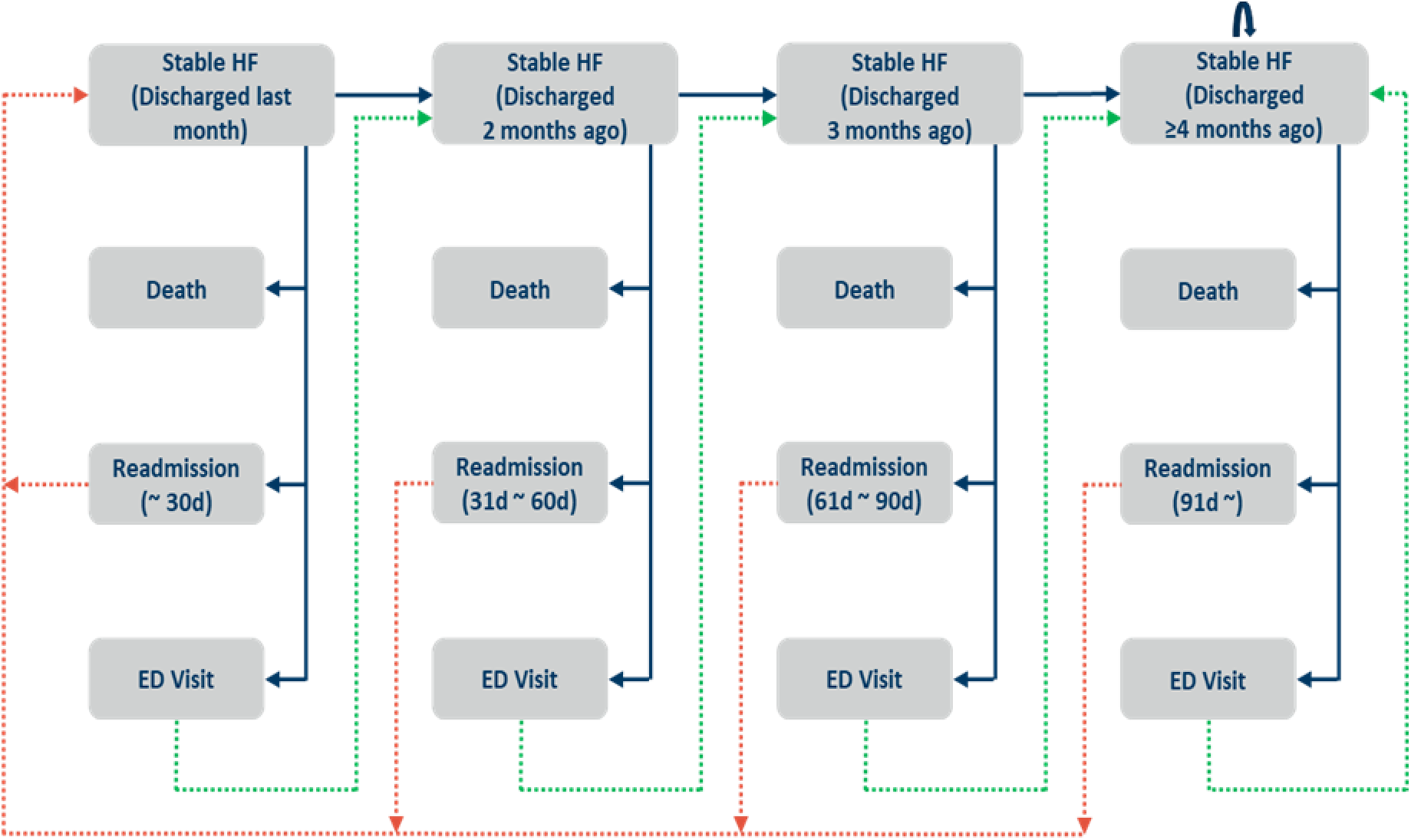
Model structure. Our model builds on previously published heart failure Markov models, taking into account the four primary health states for patients discharged from the hospital for HF: stable, death, readmission, or ED visit (blue solid and green dotted lines). Our model allows for differentiated rates of hospital readmission based on whether the patient has been discharged from the hospital within 30 days, between 31-60 days, between 61-90 days, and after 91 days (red dotted lines), since the rate of rehospitalization for HF differs significantly by the time from the last hospital discharge. This approach allows for differential time impacts after the hospitalized HF patient is discharged from the hospital. (ED = emergency department; HF = heart failure.)

The model outcomes included both health and economic outputs. Health outcomes include LYs gained and quality adjusted life-years gained. Cost captured included pharmaceutical and various types of medical costs. The economic value of sotagliflozin was measured based on incremental cost-effectiveness ratio.

### Model Inputs

Key model inputs included treatment efficacy and safety, health-related quality of life, and cost (Table 1; additional details Table S1).

**Table 1:** Table of inputs.

| Category | Item | Baseline | Source |
| --- | --- | --- | --- |
| Clinical<br>Input | Hazard Ratio<br>(Sotagliflozin vs. SoC) |  |  |
|  | Death | 0.820 | Bhatt et al. (2021) |
|  | Readmission | 0.650 | Bhatt et al. (2021) |
|  | ED Visit<sup>a</sup> | 0.600 | Bhatt et al. (2021) |
|  | Hospital readmission rate |  |  |
|  | 30-day readmission | 22.30% | Kilgore et al. (2017) |
|  | 60-day readmission | 33.30% | Kilgore et al. (2017) |
|  | 90-day readmission | 40.20% | Kilgore et al. (2017) |
|  | 1-year readmission | 43.05% | Greiner et al. (2012) |
|  | ED visit rate | 15.25% | Blecker et al. (2014), Greiner et al. (2012) |
|  | 1-year mortality rate | 36.30% | Gupta et al. (2018) |
| Utility | Stable HF | 0.800 | Alsumali et al. (2021) |
|  | Rehospitalization/ED visit | 0.723 | Alsumali et al. (2021) |
| Cost | Pharmacy cost | \$598 | Red Book |
| | Hospitalization cost | \$10,540 | CMS (2019) |
| | ED visit cost | \$3,845 | Jackson et al. (2018) |

<sup>a</sup> ED = emergency department; HF = heart failure; SoC = standard of care

#### Efficacy

Treatment efficacy was estimated from published literature on the SOLOIST-WHF trial and an internal clinical study report (CSR) of the trial ^5^. The efficacy of sotagliflozin was based on monthly rates of rehospitalization, emergency department visits, and death in each of the treatment (i.e., sotagliflozin) and control (i.e., SoC) groups. Specifically, sotagliflozin reduced the number of hospitalizations (hazard rate: 0.337 vs. 0.519 for treatment and control groups; hazard ratio (0.65), emergency department visits (0.069 vs. 0.121; 0.60), and death from any cause (0.135 vs. 0.163; 0.82) relative to SoC. To calculate the rehospitalization rates for the treatment group, we applied the hazard ratio of readmission to the 30/60/90-days and 1-year readmission rates of hospitalized heart failure patients in the U.S., which were used as the readmission rates for the control group ^14,16^. In the same way, we calculated the mortality rate of the treatment group using the hazard ratio of death and the mortality rate of the control group referenced from literature ^15^. We utilized the hazard rates of the rehospitalization and emergency department visit from CSR and the 1-year rehospitalization rate for the control group to estimate the emergency department visit rate of the control group and the hazard ratio of emergency department visit was applied to this rate to generate the emergency department visit rate of the treatment group ^13,16^. We derived the Markov transition probabilities using a numerical approach to fit the simulated distribution of the study population to the efficacy parameters under a proportional hazard assumption. The estimated monthly transition probabilities of rehospitalization, emergency department visit, and death were lower for the sotagliflozin group compared the SoC group.

#### Safety

As reported in the SOLOIST-WHF trial, adverse events included hypotension, urinary tract infection (UTI), diarrhea, pneumonia, hyperkalemia, acute kidney injury, and hypoglycemia^5^. The monthly adverse event probabilities were calculated using the median follow-up months and the adverse event rates over the follow-up periods that were identified from SOLOIST-WHF trial. The probabilities were lower for the treatment group when compared to the control group for UTI, pneumonia, hyperkalemia, and acute kidney injury, and higher for hypotension, diarrhea, and hypoglycemia.

#### Health-related Quality-of-Life

Health-related quality-of-life (HQoL) estimates—also known in economics as utilities— were estimated from two components: (i) HQoL differences between people in the stable disease, rehospitalization, and emergency department visit health states, and (ii) the impact of treatment adverse events on HQoL (Table S1). To translate the health state quality of life into utilities, we relied on baseline EQ-5D values provided from a U.S. Medicare perspective cost-effectiveness study of a soluble guanylate cyclase stimulator, vericiguat, for heart failure following a worsening heart failure event ^10^. The disutilities associated with adverse events were identified from multiple peer-reviewed sources ^17–22^.

#### Costs

Costs for both the health states and adverse events were calculated based on four categories: pharmaceutical costs, rehospitalization costs, emergency department visit costs, and adverse event costs. We assumed that all other treatment costs outside of these four categories did not change between the treatment and control arms. All costs were then inflated to 2022 USD using the Bureau of Labor Statistics Consumer Price Index Medical Care ^23^.

Incremental pharmaceutical costs by sotagliflozin were set at its wholesale acquisition cost of $598 per 30 days under an assumption of no difference in utilization of non-sotagliflozin pharmaceuticals—beyond sotagliflozin—for the hospitalized heart failure population ^24^. The baseline hospitalization costs were calculated using a Medicare payment for the Diagnosis Related Group codes 291-293 from the Centers for Medicare and Medicaid Services (CMS) ^25^. The adverse event-associated costs were all pulled from literature except for hypotension, which was calculated from the Medicare Physician Fee Schedule for 99213 of the Current Procedural Terminology (CPT) ^25^.

#### Sensitivity/Scenario Analyses

We performed a one-way sensitivity analysis using the systemic variation in the model parameters (Table S2). The lower and upper bounds, which were 10% lower and higher than the baseline values, were tested for all parameters except for treatment efficacy estimates, utility of stable health state, and the monthly cost of sotagliflozin. For the efficacy estimates, we employed the 95% confidence intervals of the hazard ratios of rehospitalization and emergency department visits reported in clinical study report of the SOLOIST-WHF trial. In the case of the hazard ratio of death, the upper bound of the 95% confidence interval was 1.23. Since it is unrealistic that the mortality risk soars by 23% by sotagliflozin use, we limited the variation of the hazard ratio of death by a 50% of the gap between the baseline hazard ratio and a hazard ratio of 1. We limited the variation of the utility of stable disease to the difference between its baseline value and the baseline utility of hospitalization/ emergency department visit. To see the model sensitivity to the pharmacy price, we used the range of the monthly cost of sotagliflozin between $498 and $698.

Additionally, a series of scenario analyses were performed. First, we examined a different rehospitalization rate from literature that assesses the mortality and rehospitalization rates of patients with heart failure associated with diabetes and depression. To match the study results, the baseline rehospitalization rate was increased by 7.8% ^26^. Second, benefits of early initiation of sotagliflozin were demonstrated from a further investigation in which the study population was limited to the patients who began study treatment on or before hospital discharge after an episode of worsening heart failure: the 30- and 90-day rehospitalization hazard ratios were 0.48 and 0.52, respectively ^27^. The updated hazard ratios were applied under the second scenario analysis. Third, while our baseline approach used a 3% discount rate, we also recalculated the results without discounting future health benefits or costs. Last, we examined how the treatment value—as measured by the incremental cost effectiveness ratio—changes when the study time horizon changes. The analyses look at whether sotagliflozin provides value to society in the short-run (e.g., within 1, 2, and 3 years) as compared to only value in the long run (e.g., over the full 30-year period).

This study does not involve human subjects or include any access to identifiable private information.

## Results

### Base Case

Mirroring the results of the SOLOIST-WHF trial, the model predicted that sotagliflozin would reduce hospital readmissions and emergency department visits relative to SoC. Model dynamics showed that a majority of patients in both arms died prior to the 10-year period, however the proportion of patients in the control group without heart failure recurrence (red solid lines) was higher than that in the control group (red dotted line) (Figure 2). Meanwhile, the proportions of rehospitalization (blue lines) and emergency department visits (green lines) were lower in the treatment group (solid blue and green lines). The efficacy of sotagliflozin is also supported by the more modest decline of the survival curve of the treatment group (orange line) compared to the control group (dark blue line) (Figure S1). These results are consistent with life-years gained of 0.60 years by the treatment group (3.36 vs. 2.76 for the treatment and control groups).

**Figure 2:**
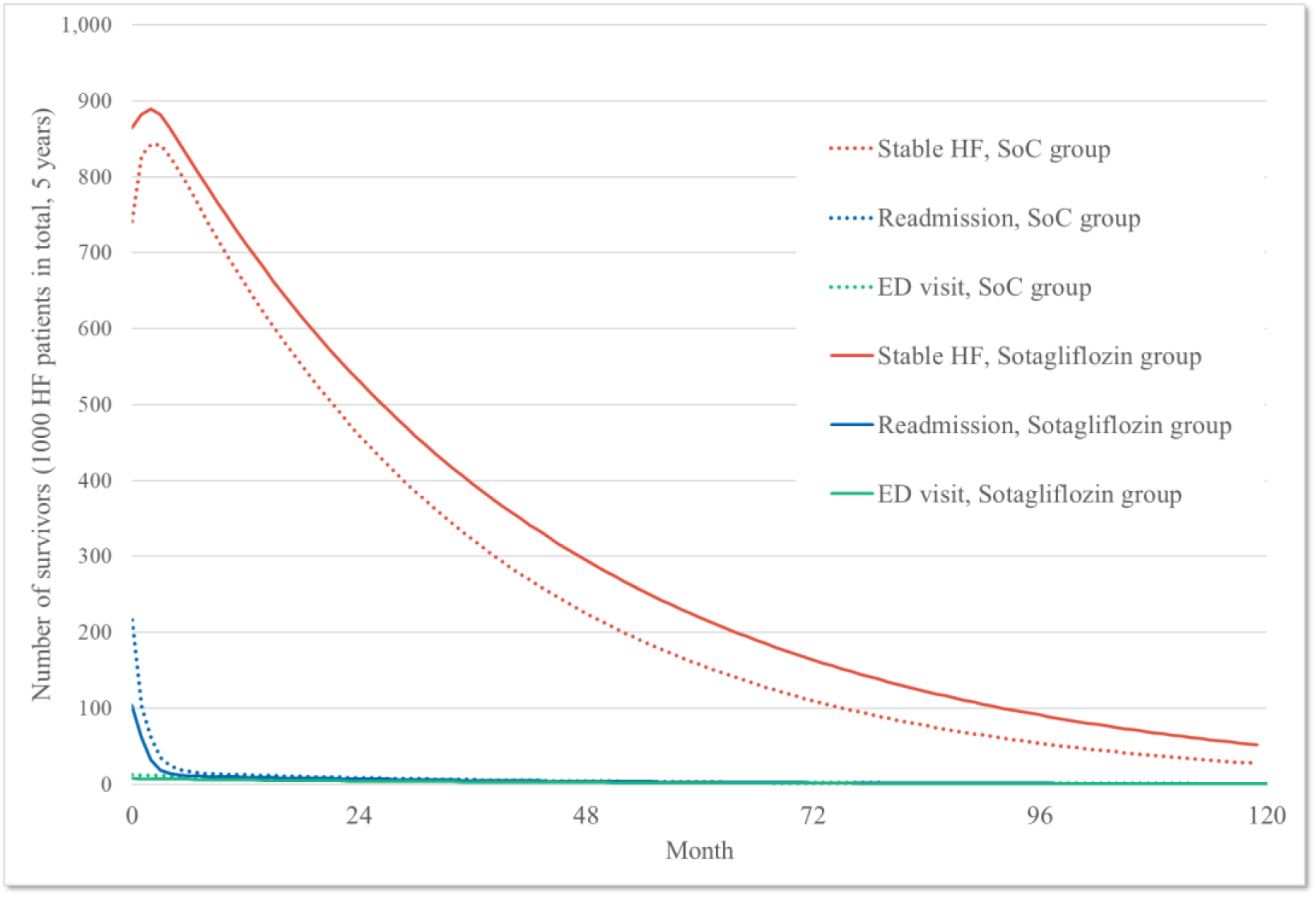
Dynamics of distribution of heart failure patients after initial hospital discharge, sotagliflozin vs. standard of care groups. According to the Markov model, sotagliflozin is projected to decrease hospital readmissions and ED visits compared to the SoC. Although a significant number of patients in both groups died before the 10-year period, the treatment group (represented by red solid lines) had a higher proportion of patients without HF recurrence compared to the control group (represented by red dotted line). On the other hand, the treatment group (solid blue and green lines) showed lower proportions of rehospitalization and ED visits in comparison to the control group (dotted blue and green lines). (ED = emergency department; HF = heart failure; SoC = standard of care.)

Over the 30-year time horizon, use of sotagliflozin significantly reduced the risk of hospital readmissions, emergency department visits, and deaths in the observed cohort (Figure 3). The annualized rehospitalization and emergency department visit rates estimated over the study period for the treatment group were lower than the control group by 34.5% (0.228 vs. 0.348 per year, difference: -0.120) and 40.0% (0.091 vs. 0.153 per year, difference: -0.061) respectively. The annualized mortality rate of the treatment group decreased by 18.0% compared to the control group (0.298 vs. 0.363, difference: -0.065). The risk reduction of the heart failure recurrence requiring rehospitalization by sotagliflozin is the driving factor of the better health outcomes in the treatment group.

**Figure 3:**
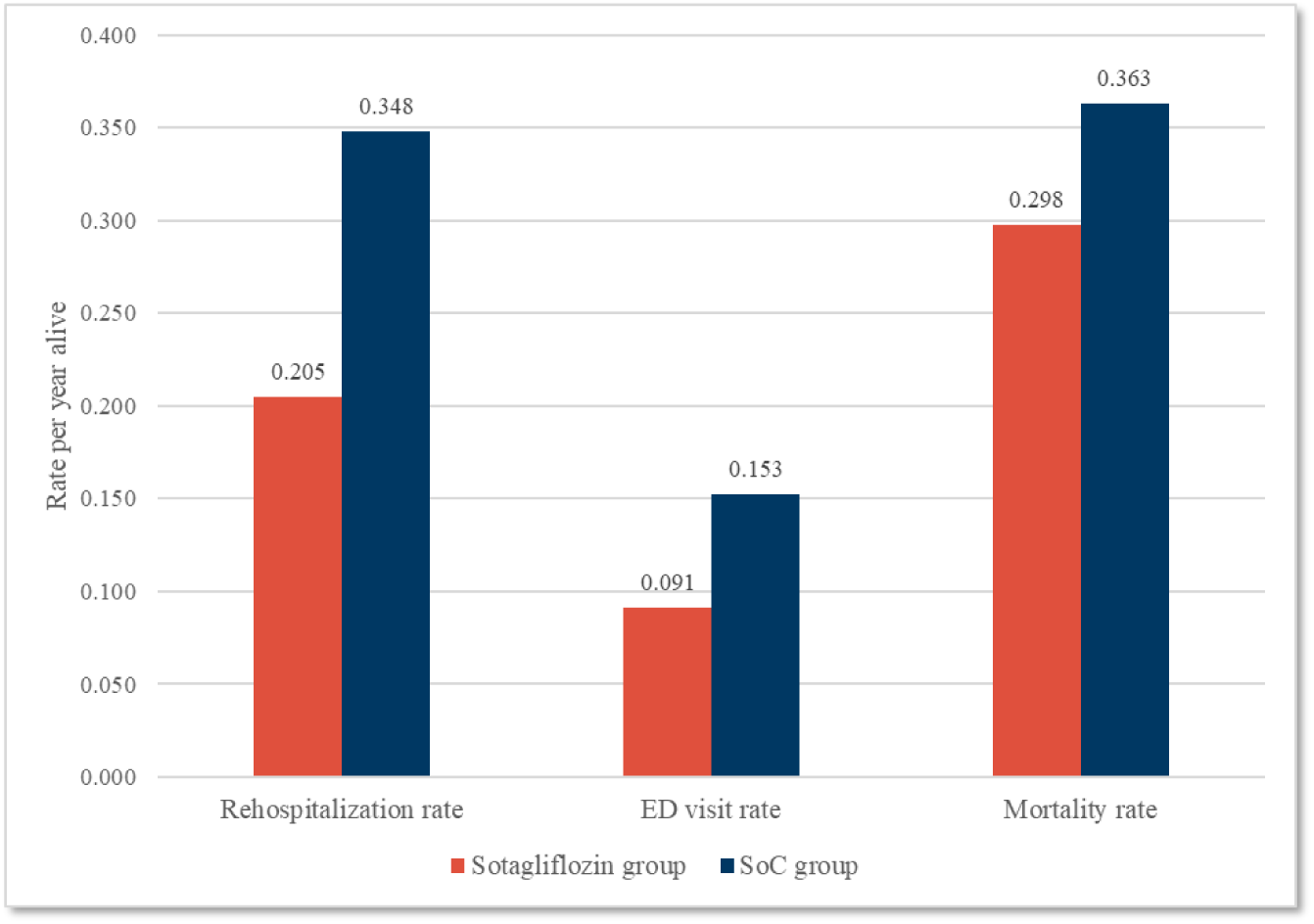
Health outcomes comparison, sotagliflozin vs. standard of care groups. As per the Markov model, the treatment group exhibited significantly lower annualized rehospitalization and ED visit rates over the study period compared to the control group, with reductions of 34.5% (0.228 vs. 0.348 per year, difference = -0.120) and 40.0% (0.091 vs. 0.153 per year, difference = -0.061), respectively. Additionally, the treatment group’s annualized mortality rate decreased by 18.0% in comparison to the control group (0.298 vs. 0.363, difference = -0.065). (ED = emergency department; SoC = standard of care.)

Due to reduced mortality and improved quality of life produced by fewer hospital readmissions and emergency department visits, quality adjusted life-years were estimated to be higher among patients in the treatment group compared to the control group (Table 2, Figure S1). Discounted quality adjusted life-years gained over 30 years was 0.425 (2.305 vs. 1.880). The improvement was due to patients in the treatment group spending more time in the “Stable heart failure” health state due to reductions in mortality, rehospitalizations, and emergency department visits. Although the loss of quality adjusted life-years by rehospitalizations and emergency department visits partly offset the quality adjusted life-years gained during the period with the stable health state (“Stable heart failure”), the magnitude of the quality adjusted life-years change in the stable health state (2.249 vs. 1.808, difference: 0.441) overwhelmed the decrement of quality adjusted life-years in the other two events (0.056 vs. 0.072, difference: -0.016).

**Table 2:**
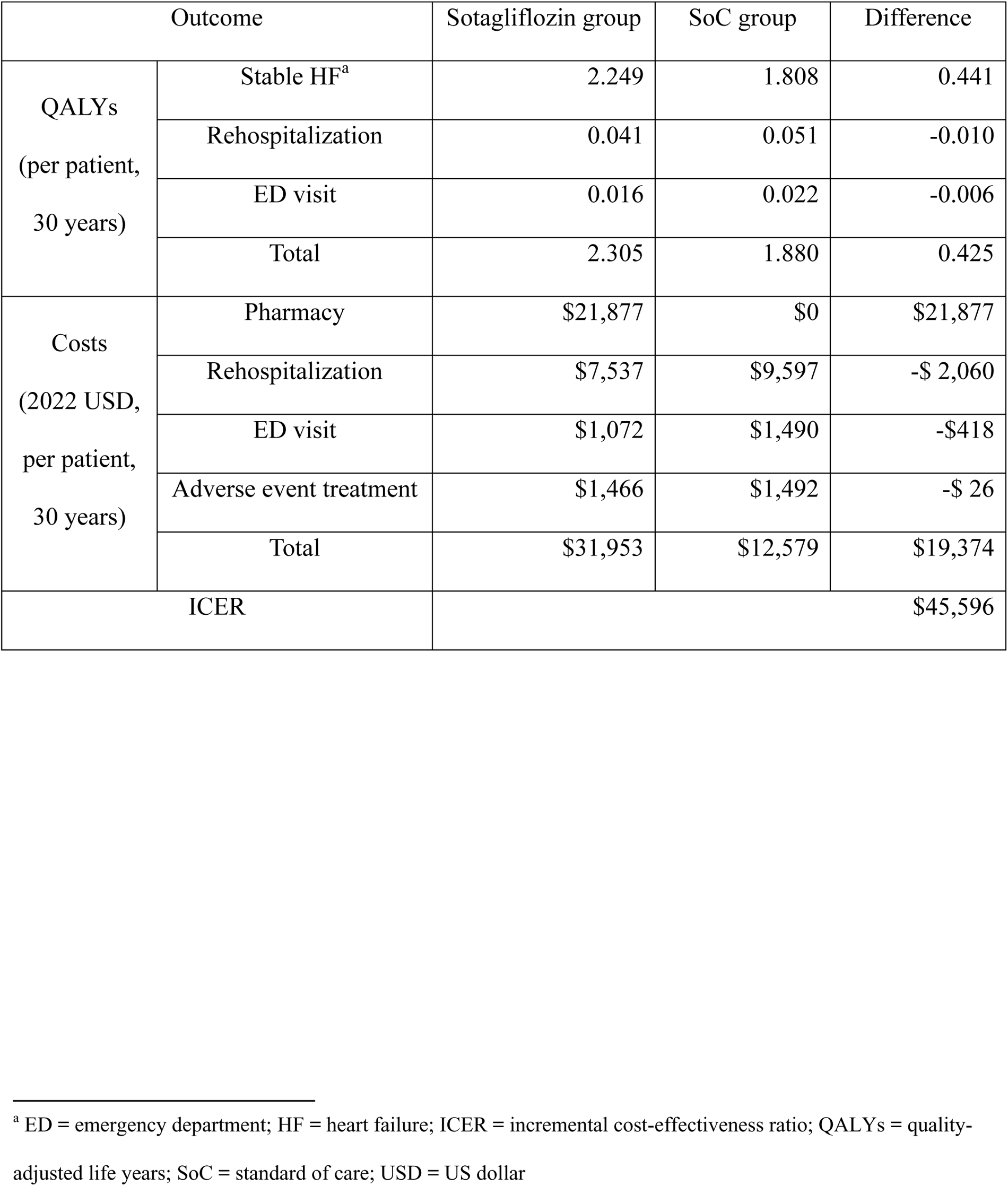
Summary of cost effectiveness analysis results, base case.

The discounted cost of care of the treatment group over 30 years was about 2.5-fold of that of the control group ($31,953 vs. $12,579, difference: $19,374) (Table 2, Figure 4). About three quarters of the total costs were explained by the pharmacy cost for the treatment group ($21,877, 68.5%) and by the rehospitalization cost for the control group ($9,597, 76.3%). Given that magnitudes of the cost reduction by reduced rehospitalizations, emergency department visits, and adverse events of the treatment group ($7,537 vs. $9,597 for rehospitalizations, difference: – $2,060; $1,072 vs. $1,490, -$418 for emergency department visits; $1,466 vs. $1,492, -$26 for adverse events treatment) are relatively small compared to the cost increment by sotagliflozin ($21,877), the pharmacy cost plays a pivotal role in determining the opportunity costs of sotagliflozin use.

**Figure 4:**
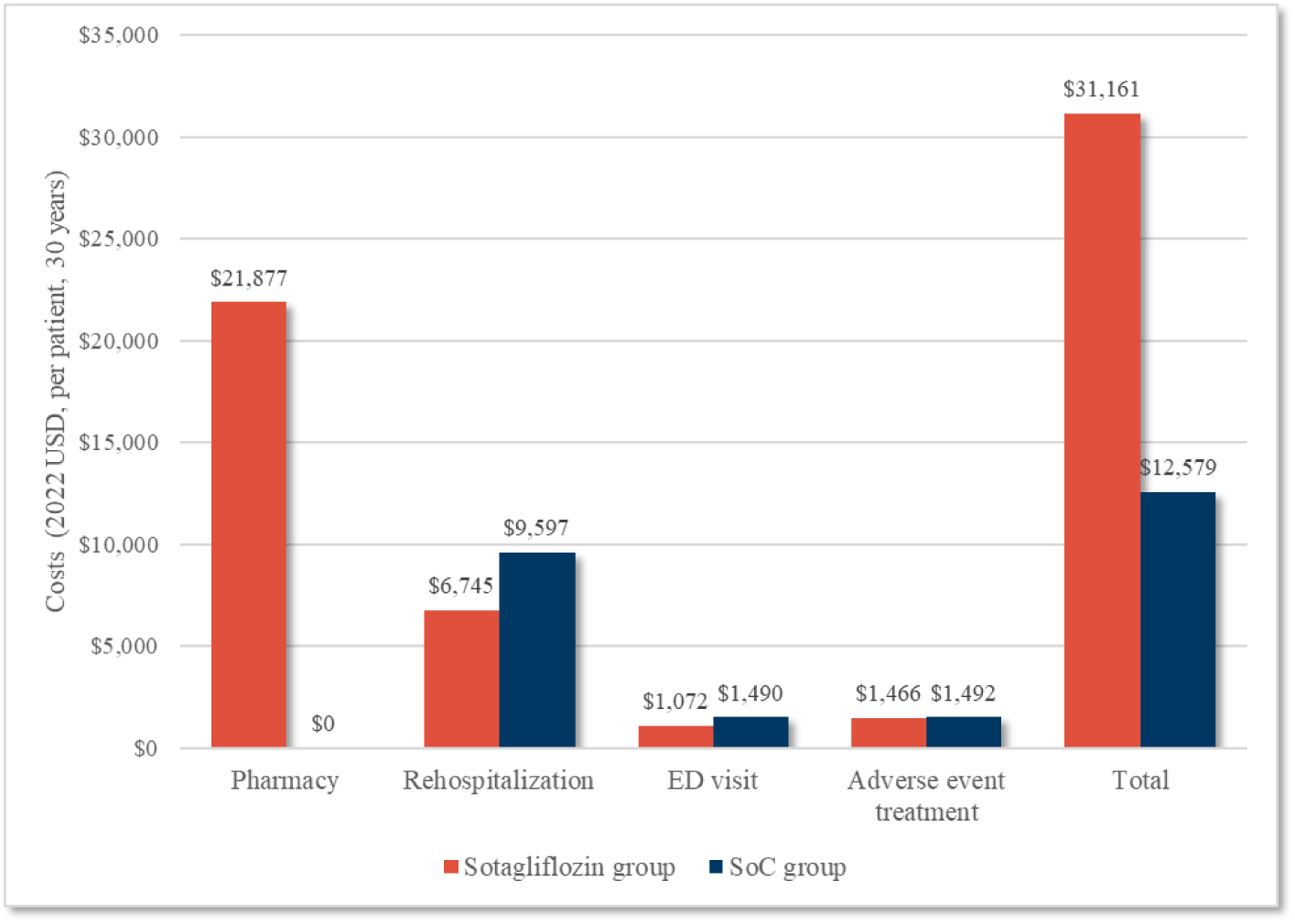
Cost comparison, sotagliflozin vs. standard of care groups. The discounted cost of care of the treatment group over 30 years was about 2.5-fold of that of the control group ($31,953 vs. $12,579). Nearly three quarters of the total costs for the treatment group were explained by the pharmacy cost ($21,877, 68.5%) and by the rehospitalization cost for the control group ($9,597, 76.3%). Cost reductions are relatively small for rehospitalizations, ED visits, and adverse events in the treatment group (-$2,060, -$418, and – $26) compared to the increase in pharmacy cost. (ED = emergency department; SoC = standard of care; USD = US dollar.)

Sotagliflozin is a cost-effective addition to SoC for hospitalized heart failure patients (Table 2). At a 3% discount rate, incremental cost-effectiveness ratio is $45,596 per quality adjusted life-year gained, which is lower than the commonly used U.S. payer willingness-to-pay (WTP) per quality adjusted life-year of $100,000 ^28^.

### Sensitivity/Scenario Analyses

The results of the one-way sensitivity analysis presented in the tornado diagram in Figure S2 indicate that sotagliflozin is cost-effective compared to SoC for any possible variation of parameters. The model was shown to be sensitive to the efficacy of sotagliflozin, such as the hazard ratios of death, rehospitalization, and emergency department visit (range of incremental cost-effectiveness ratio: [$33,246, $77,525], [$41,367, $51,215], [$44,422, $47,685]). The pharmaceutical cost and the utility in the stable health state were also important factors in determining the economic value of sotagliflozin ([$36,986, $54,207], [$41,440, $50,680]).

Under the scenarios in which higher rehospitalization rates provided from literature were considered, treatment was initiated on or prior to hospital discharge, or benefits and costs were not discounted, the results did not make a significant difference in the economic value of sotagliflozin compared to the baseline (incremental cost-effectiveness ratio: $45,298, $43,684, $43,143) (Figure S2, Central Illustration). Additionally, sotagliflozin was cost-effective in the short run as well as in the long run: incremental cost-effectiveness ratio was $93,079 and $77,710 for the 2- and 3-year models, respectively. However, incremental cost-effectiveness ratio exceeded the cost-effectiveness threshold in the 1-year model ($114,985) because the study period was too short to have the model consider the benefit of lower mortality rate of sotagliflozin.

## Discussion

Using a novel, first-order Markov chain structure that allows for rehospitalization rates to vary by time from discharge, we found that sotagliflozin was cost-effective for heart failure treatment among patients with both worsening heart failure and comorbid diabetes. Specifically, the estimated incremental cost-effectiveness ratio from sotagliflozin use was $45,596 per quality adjusted life-year gained. Considering the model’s sensitivity to the cost of sotagliflozin and that the model implements a wholesale acquisition cost for sotagliflozin, it is reasonable to assume that the estimated incremental cost-effectiveness ratio is conservative relative to real-world costs that payers may negotiate. Over the 30-year time horizon, sotagliflozin resulted in reduced risk of hospital readmissions, emergency department visits, and deaths, effectively increasing the quality adjusted life-years in the observed cohort. Although the overall cost of care increased, largely due to increased pharmacy costs, this was offset by fewer costs associated with lower rates of hospital readmissions and emergency department visits. Overall, the results were robust to a variety of sensitivity and scenario analyses.

As the first CEA of sotagliflozin, our study provides a number of unique contributions to the existing literature. While there are several different heart failure models, our model allows the flexibility to have different rehospitalization rates dynamically over time as readmission rates are higher within 30 days as compared to 31-60 days, 61-90 days or after 90 days. Other heart failure CEA models that have studied SGLT2 inhibitors, such as empagliflozin and dapagliflozin, have found them to be cost-effective additions to SoC, albeit at differing willingness-to-pay thresholds ^7,8^.

This study has several limitations. First, this study was conducted based on the SOLOIST-WHF trial with a short follow up period. As a result, the long-term benefits had to be extrapolated based on the available data, and therefore, the total benefits in the results are highly dependent on the chosen parameters in those calculations. However, this study shows that the treatment is largely cost-effective even in the first year. Second, this study uses health state utilities rather than the utilities estimated in the SOLIST-WHF trial. Third, due to lack of reliable data, the model does not consider rebates of the pharmacy costs, further limiting the results of the study. The inclusion of rebates in the model would improve the economic value of sotagliflozin from the payer perspective. Fourth, non-adherence is not included in the model due to the difficulty in obtaining accurate adherence levels. Further, the effect of nonadherence on cost-effectiveness is unclear due to competing effects. Lastly, the model assumes consistent Medicare prices across all patients. Commercial patients would have a higher cost-effectiveness because they pay higher costs for hospitalization and emergency department visits ^29^.

Despite these limitations, this study found that sotagliflozin is a cost-effective treatment for heart failure among patients with type 2 diabetes and a recent heart failure hospitalization or urgent care visit. The estimated incremental cost effectiveness ratio of approximately $46,000 per quality adjusted life-year represent high value at commonly used willingness to pay thresholds in the US.

## Data Availability

The data that support the findings of this study are available are publicly available and can be found within the references of the manuscript.

## Abbreviation list

CEA: Cost-effectiveness analysis
CMS: Centers for Medicare and Medicaid Services
CPT: Current Procedural Terminology
CSR: Clinical study report
HQoL: Health-related quality-of-life
LY: Life-year
SGLT1: Sodium-glucose cotransporter type 1
SGLT2: Sodium-glucose cotransporter type 2
SoC: Standard of care
UTI: Urinary tract infection

## Acknowledgments

We would like to thank Natalia Vasquez for their contribution towards the manuscript draft.

---

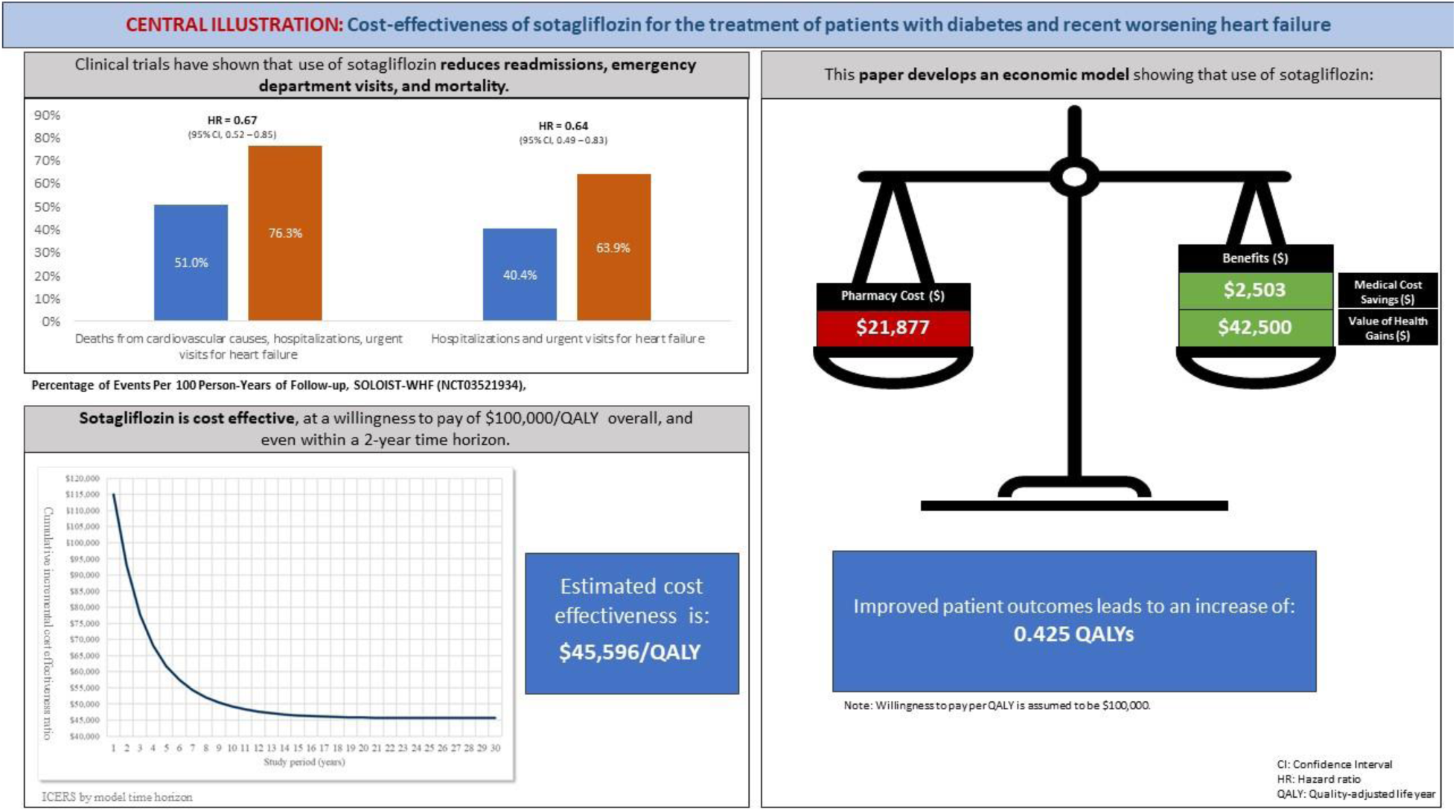
Central Illustration. Evidence from clinical trials show that sotagliflozin reduced readmissions, emergency department visits (HR = 0.64), and deaths (HR = 0.67) compared to SoC among patients with diabetes and recent worsening HF. Sotagliflozin also reduced occurrence of the majority of common adverse events. This economic model estimated that use of sotagliflozin increased costs by $19,374 and resulted in a net gain in QALYs of 0.425. Estimated ICER was $45, 596 per QALY. (HR = hazard ratio; ICER = incremental cost-effectiveness ratio; QALYs = quality-adjusted life years; SoC = standard of care.)

